# Women are Prone to Age-Related aortic Stiffness

**DOI:** 10.1101/2023.05.25.23290560

**Authors:** Zixuan Meng, LeLe Cheng, Wenjun Liu, Yue Yu, Hui Liu, Guolin Yao, Jian Yang, Yue Wu, Zhijie Jian

## Abstract

**Aim:** To investigate sex differences in the aortic aging by analyzing aortic diameter and tortuosity in different segments of aorta across the age spectrum, using enhanced CT imaging.

**Method:** Between July 2021 and April 2022, a retrospective study screened patients with chest and abdomen contrast CT images. The outer edge-to-outer edge method was utilized to measure aortic diameters at five aortic levels, while arterial tortuosity of various segments was measured and calculated using imaging software. Mean values were compared at different age groups, including by sex, and correlation with age was determined. To validate the coherence of arterial elasticity and anticipated age-related arterial alterations, a subset of data from a previously published article in BMJ Open was extracted for the purpose of examining the correlation between age and arterial stiffness, stratified by sexes.

**Results:** 208 participants (56.6% men, mean age 60.13±16.33 years old, mean BMI 23.07±4.03 kg/m2, mean BSA 1.70±0.19 m2) were enrolled in this study. The BSA-adjusted aortic diameters showed a positive correlation with age in both sexes, but females demonstrated a more rapid increase in progressive aortic diameters throughout their lifespan than males. In the age groups of 60-69 and above 80 years old, males exhibited significantly larger L1 measurements than females. Conversely, in individuals over 80 years old, females displayed greater L3 values compared to their male counterparts. However, no sexual disparities were observed for L2, L4 and L5 across all ages. Females exhibited greater aortic tortuosity in the descending thoracic region compared to males across all age groups, whereas this sex-based distribution of aortic and abdominal tortuosity was only evident among individuals over 40 years old. The tortuosity of the aorta and descending thoracic aorta exhibits a marked increase with advancing age, particularly in females, while a non-significant linear correlation is observed between abdominal aortic tortuosity and age in both genders. BaPWV consistently increased with age in both males and females, but the increase was more significant in females. Although males initially had higher arterial stiffness, females surpassed them as they aged.

**Conclusion:** Patterns of vascular aging in aortic morphology differ between the sexes across the life course, with women experiencing more significant changes, especially in advanced age groups.

## Introduction

Cardiovascular diseases (CVD) have been a momentous health issue globally. It is well known that cardiovascular disease mortality increases exponentially with age in both males and females, and is higher in older women(1, 2). In addition, CVD exhibits sex-specific differences with women experiencing a rapid increase in cardiovascular risk and atypical symptoms after menopause(3, 4). This discrepancy may be attributed to higher prevalence of cardiovascular risk factors (e.g., obesity, diabetes) in older women than men in the same age group(5). Compared to these pathological conditions, the understanding of physiological vascular aging as a crucial factor in the development of CVD is less comprehensive(6), and sex differences in the aging process may explain why certain sex are more susceptible to CVD risk factors.

Aging causes changes in the morphology of the aorta, leading to tortuosity, dilation, and stiffness(7, 8), which affect the hemodynamics of the aorta, resulting in an increase in cardiac afterload, over-stretching of the aortic wall, and reducing coronary perfusion(9, 10). Eventually, these morphological changes in the aorta could exacerbate atherosclerosis, increased the incidence of ischemic events such as stroke, and aneurysms(11, 12). The progressive age-related changes in aortic anatomy can be measured with many imaging modalities, confirming the existence of significant sex differences in aortic diameter and tortuosity by using enhanced computed tomography (CT) and magnetic resonance imaging (MRI)(13, 14). A cross-sectional study from the Multi-Ethnic Study of Atherosclerosis (MESA), using aorta MRI, had reported that the ascending thoracic aorta diameter increased gradually with aging for both sexes, while after adjusting for body surface area (BSA), women consistently exhibited greater normal values than men across all ages(14).The investigation of age-related changes in the descending aorta using enhanced CT revealed that the tortuosity increases more significantly with age in women than in men(15). However, these studies have only focused on the certain segment of the aorta, targeting either diameter or tortuosity. The role of sex in the aging of overall aortic morphology remains largely unexamined. Therefore, further investigation should be required to explore sex differences in evolutionary trends throughout the lifespan.

Our study aimed to investigate sex differences in the aortic aging by analyzing aortic diameter and tortuosity in different segments of aorta across the age spectrum, using enhanced CT imaging.

Additionally, we verified our results by examining the relationship between arterial stiffness and age from BMJ Open data to gain a comprehensive understanding of adults aortic aging for both sexes.

## Materials and Methods

### Study population and data collection

A retrospective study was conducted at the First Affiliated Hospital of Xi’an Jiaotong University, focusing on patients who underwent chest and abdomen contrast CT imaging between July 2021 and April 2022. The study excluded patients who had previously been diagnosed with hypertension, diabetes, or kidney disease, as well as those who had aneurysms, dissection, vascular malformation, or variation on CT scan. Inadequate quality CT scans with significant artifacts were also excluded. Finally, individuals with insufficient clinical data for analysis were excluded from the study. Multiple demographics were obtained from the medical history records, including age, sex, weight, height, and body mass index (BMI). Meanwhile, we standardized the aorta diameter and tortuosity by BSA to exclude the effect of diverse body shapes on the data analysis. BSA was calculated by Dubois formula 1 which was referred as BSA (m^2^) = 0.007184 × [height (cm)^(0.725)^] × [height(kg)^(0.425)^]. Current smoking was defined as positive smoking, Related laboratory data including triglycerides, cholesterol, low-density lipoprotein cholesterol (LDL), high-density lipoprotein cholesterol (HDL) and serum glucose were assayed by automated chemistry analyzer. All the laboratory tests were conducted 48 hours before the CT scan. Written informed consent was obtained from all study participants, and this study was approved by the ethic committee approval of the First Affiliated Hospital of Xi’an Jiaotong University (Ethical approval number: XJTU1AF2020LSL-018).

We also used partial data from a published article in BMJ Open, an open-access scientific journal, named “Association between serum γ-glutamyltranspeptidase and atherosclerosis: a population-based cross-sectional study” which provides access to the original data(16). This cross-sectional study was performed at the Medical Health Checkup Center of Murakami Memorial Hospital, Gifu city, Japan from March 2004 to December 2012. 1445 participants receiving a medical health check-up program including pulse wave velocity and abdominal ultrasonography were screened. The patients’ personal information in the original data is anonymous, therefore informed consent was not required. Data such as age, sex, and branchial ankle pulse wave velocity (baPWV) are extracted from this article to analyze the sex disparities in the association between age and baPWV.

### Assessment of aortic diameters and tortuosity index calculation

Two 256-slice CT scanners, namely the Philips Brilliance iCT from Medical Systems in Best, The Netherlands, and the Revolution CT from GE Healthcare in Milwaukee, WI, were utilized to conduct a thoracoabdominal enhanced CT scan. The scan encompassed the area from the lung apices to the iliac crest and was performed without ECG gating. The data were subjected to analysis using a standard post-processing workstation (uInnovation-CT, R001, United Imaging Healthcare, Shanghai, China) equipped with commercially available volume viewer software. The aortic diameter was measured at five distinct levels, namely the ascending aorta (L1), situated 1cm distal to the sinotubular junction; the descending aorta (L2), aligned with L1; the intersection of aorta and diaphragm (L3); 3cm above the level of the aortic bifurcation (L5), and the midpoint between L3 and L5 (L4), using the outer edge-to-outer edge method (Fig 1A). Next, we calculated the tortuosity of the whole aorta, thoracic aorta, and abdominal aorta separately. Subsequently, the tortuosity of the entire aorta, thoracic aorta, and abdominal aorta was computed individually. A proficient radiologist specializing in cardiovascular imaging accurately identified the aorta of the corresponding segment by pinpointing two points on the corresponding anatomical landmark. As an illustration, the measurement of aortic tortuosity was conducted on the aorta, which was defined as commencing at the aortic valve and terminating at the bifurcation of the iliac artery. Subsequently, an automated centerline was generated between the two points (Fig 1B). To mitigate the impact of varying body shapes on data analysis, we standardized the aorta diameter and tortuosity by BSA.

**Figure 1.**
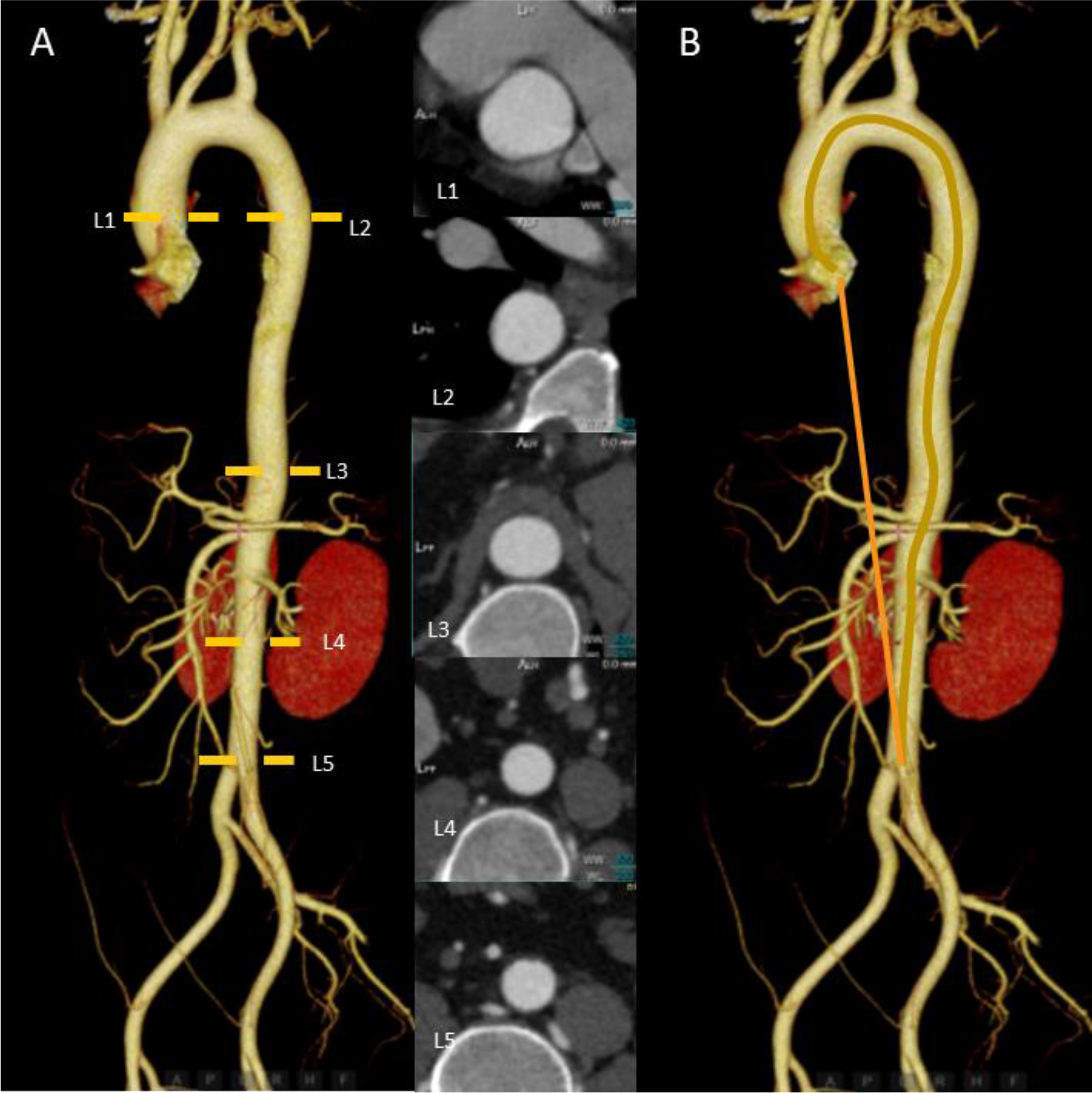
The measurements of aortic morphology. A, Sites of diameter measurements of the aorta; B, Measurement of tortuosity index. The blue line is the true length measurement of the vessel, and the orange line is the straight-line length. The tortuosity index of the aorta is the true length / the straight-line length.

### Statistical analysis

Patients enrolled are divided into different age groups: <40, 40∼49, 50∼59,60∼69,70∼79, ≥ 80. All the continuous data in accordance with normality distribution are presented as mean ± SD and compared by t-test between groups, the Kruskal-Wallis test is used to compare continuous data if not normally distributed. The categorical variables are presented as numbers and percentages, compared by χ^2^ test. We also used multiple linear regression analysis to analyze the potential factors associated with aorta aging. The association of BSA-indexed aortic dimensions with aging was assessed by linear regression models in each sex. A *p*-value < 0.05 was regarded as statistically significant for all statistical tests. All statistical analyses were performed by R software (version 4.0.2).

## Results

### Characteristics of participants

A total of 208 participants were enrolled in this study. The average age was 60.13±16.33 years old, and 56.6% of participants were men. Participants had a mean BMI of 23.07±4.03 kg/m^2^ and BSA of 1.70±0.19 m^2^ (Table 1). Age was similar in females compared with males (females 60.06±16.97 years; males 60.18±15.95 years; *p*=0.462).

**Table 1.** Characteristics of participants by sex.

| Variables | All participants<br>(N=208) | Male<br>(n=124, 56.6%) | Female<br>(n=84, 43.4%) | *p* value |
| --- | --- | --- | --- | --- |
| Age, years | 60.13±16.33 | 60.18±15.95 | 60.06±16.97 | 0.959 |
| BMI, kg/m<sup>2</sup> | 22.86(20.59,25.39) | 23.61±4.11 | 22.27±3.80 | 0.019 |
| BSA, m<sup>2</sup> | 1.71±0.19 | 1.79±0.16 | 1.57±0.16 | < 0.001 |
| Current smoking | 76 (36.5%) | 75 (60.5%) | 1 (1.2%) | < 0.001 |
| SBP, mmHg | 122(112,134.75) | 124.52±12.49 | 123.52±18.57 | 0.549 |
| DBP, mmHg | 75(70,80) | 76.55±9.02 | 74.08±9.40 | 0.058 |
| White blood cells,<br>10<sup>9</sup>/L | 5.82(4.64,7.17) | 5.96(4.64,7.4) | 5.50(4.54,7.16) | 0.548 |
| Cholesterol,<br>mmol/L | 3.97(3.28,4.70) | 4.18±1.37 | 3.83(3.18,4.54) | 0.133 |
| Albumin, g/L | 36.5(33,39.3) | 36.71±4.94 | 36.55(31.05,39.10) | 0.151 |
| Total bilirubin | 8.9(6.4,13.1) | 9.40(6.60,14.05) | 8.55(6.25,12.15) | 0.132 |
| Indirect bilirubin | 6.5(4.5,10) | 7.10(4.45,10.35) | 6.35(4.70,9.70) | 0.467 |
| Glucose | 4.65(4.12,5.39) | 5.22(4.39,6.05) | 4.77(4.29,5.70) | 0.132 |
| Urea | 4.91(4.2,5.94) | 5.58(4.58,6.85) | 4.84±1.70 | < 0.001 |
| Creatinine | 56(49,66) | 62.50(54,72) | 50.15±11.73 | < 0.001 |
| eGFR | 101.98±21.15 | 102.87±20.68 | 100.85±21.90 | 0.623 |

BMI: body mass index; BSA: body surface area; SBP: systolic blood pressure; DBP: diastolic blood pressure; CRP: C-reactive protein; eGFR: estimated glomerular filtration rate.

BMI, BSA, blood pressure, glucose, cholesterol and estimated glomerular filtration rate (eGFR) showed no significant differences between males and females. On the contrary, males exhibited significantly higher levels of urea and creatinine compared to females. The proportion of male smokers was also significantly higher than that of female smokers. The clinical characteristics are summarized in Table 1.

### Arterial diameters increased more with age in females than males

BSA-adjusted aortic diameters at all different levels demonstrated a positive correlation with age in both sexes, indicating that females exhibited a more repaid increase in progressive aortic diameters with aging throughout the life course compared to males (Fig 2). However, at the ages of 60-69 and above 80 years old, L1 was significantly larger in males than in females. The L3 of females over 80 years old was larger than that of males, whereas there were no sexual disparities observed for L2, L4 and L5 across all ages (Supplementary figure 1, Table 2).

**Figure 2.**
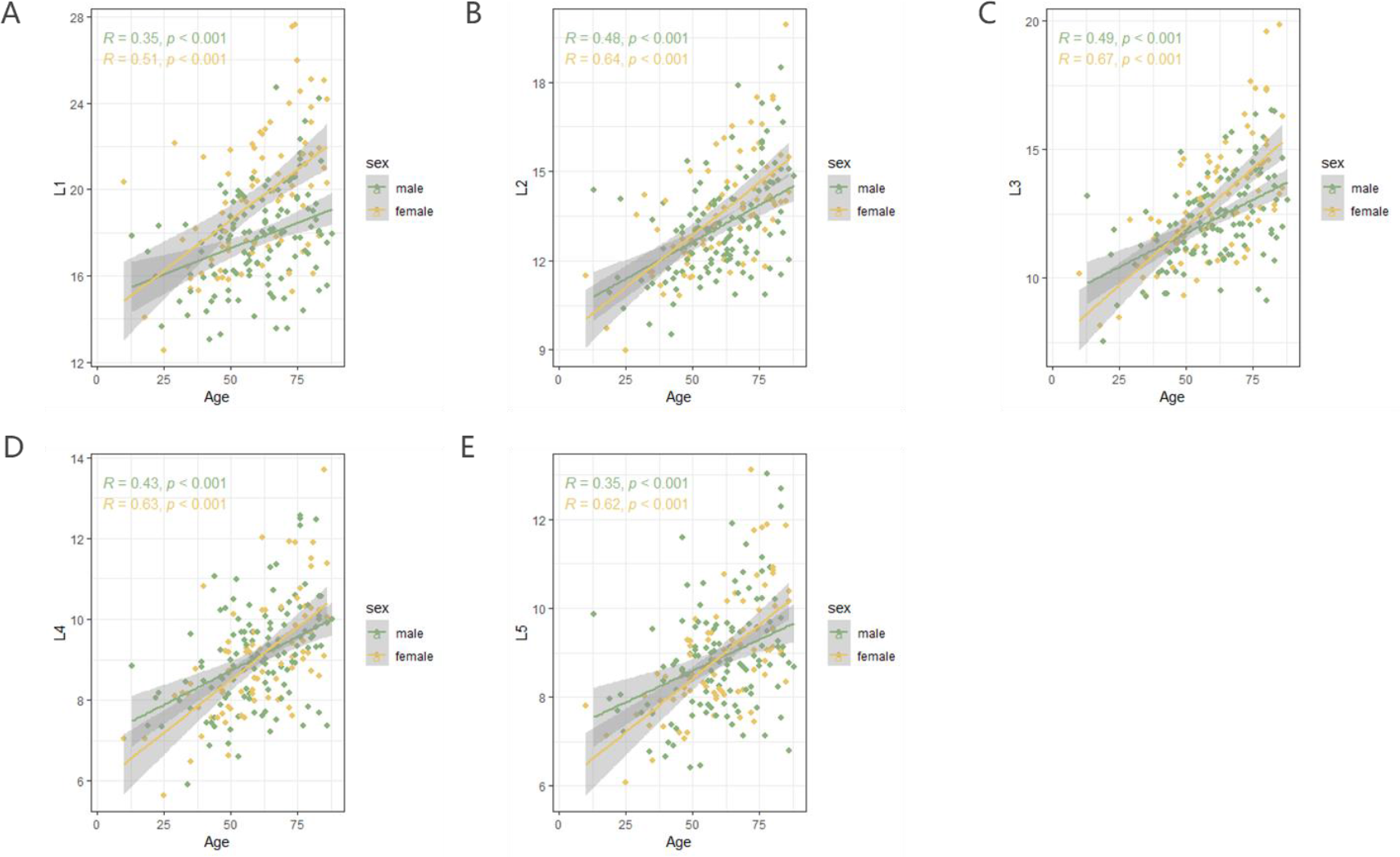
The relationship between BSA-corrected aortic diameters and age according to sex. Linear-regression analysis showed a linear relationship between age and L1(A), L2 (B), L3 (C), L4(D), L5(E) in males and females.

**Table 2.** Sex disparities in BSA-adjusted aortic diameters among different age **groups.**

| BSA-adjusted |  |  |  |  |
| --- | --- | --- | --- | --- |
| aortic | Age groups | Male | Female | *p* value |
| diameters |  |  |  |  |
| L1 | <40 | 16.35±1.60 | 16.73±2.74 | 0.928 |
|  | 40-49 | 16.40±2.06 | 17.75±1.79 | 0.116 |
|  | 50-59 | 17.86±1.62 | 19.08±1.99 | 0.069 |
|  | 60-69 | 17.94±2.17 | 20.23±2.16 | 0.003 |
|  | 70-79 | 18.85±2.52 | 21.28±3.87 | 0.054 |
|  | ≥80 | 18.39±2.27 | 21.68±2.80 | 0.007 |
| L2 | <40 | 12.03±1.55 | 11.61±1.54 | 0.833 |
|  | 40-49 | 12.12±1.28 | 12.42±0.28 | 0.561 |
|  | 50-59 | 12.85±1.07 | 13.28±1.04 | 0.281 |
|  | 60-69 | 13.15±1.48 | 13.96±1.53 | 0.095 |
|  | 70-79 | 13.90±1.69 | 14.54±1.64 | 0.267 |
|  | ≥80 | 14.66±1.93 | 15.36±1.98 | 0.462 |
| L3 | <40 | 10.72±1.64 | 10.32±1.37 | 0.487 |
|  | 40-49 | 11.13±1.35 | 11.74±1.41 | 0.125 |
|  | 50-59 | 12.24±0.93 | 12.51±1.35 | 0.377 |
|  | 60-69 | 12.44±1.47 | 12.93±1.54 | 0.384 |
|  | 70-79 | 13.36±1.64 | 14.15±2.05 | 0.292 |
|  | ≥80 | 13.01±2.07 | 15.65±2.48 | 0.014 |
| L4 | <40 | 8.10±0.97 | 7.51±0.94 | 0.134 |
|  | 40-49 | 8.38±1.17 | 8.33±1.06 | 0.955 |
|  | 50-59 | 8.97±1.02 | 8.67±0.77 | 0.257 |
|  | 60-69 | 9.13±1.05 | 9.28±1.10 | 0.811 |
|  | 70-79 | 9.74±1.36 | 9.68±1.23 | 0.989 |
|  | ≥80 | 9.73±1.22 | 10.53±1.49 | 0.193 |
| L5 | <40 | 8.10±0.90 | 7.46±0.68 | 0.091 |
|  | 40-49 | 8.38±1.34 | 8.26±0.74 | 0.750 |
|  | 50-59 | 8.71±0.92 | 8.64±0.71 | 0.779 |
|  | 60-69 | 8.88±1.10 | 8.99±1.02 | 0.772 |
|  | 70-79 | 9.57±1.37 | 9.84±1.64 | 0.692 |
|  | ≥80 | 9.35±1.59 | 10.00±1.02 | 0.145 |

### More remarkable age-related increase of segmental aortic tortuosity in females

Regarding the BSA-corrected aortic tortuosity index, a statistically significant difference was observed between genders across the majority of age groups. (Table 3). Among all age groups, females exhibited greater tortuosity in the descending thoracic aorta than males. Yet this sex distribution in the aorta and abdominal aorta appeared in people over 40 years (Supplementary figure 2). As individuals age, there is a noticeable increase in the tortuosity of the aorta and descending thoracic aorta, especially in females. However, there is no significant linear correlation between abdominal aortic tortuosity and age in either sex (Fig 3).

**Figure 3.**
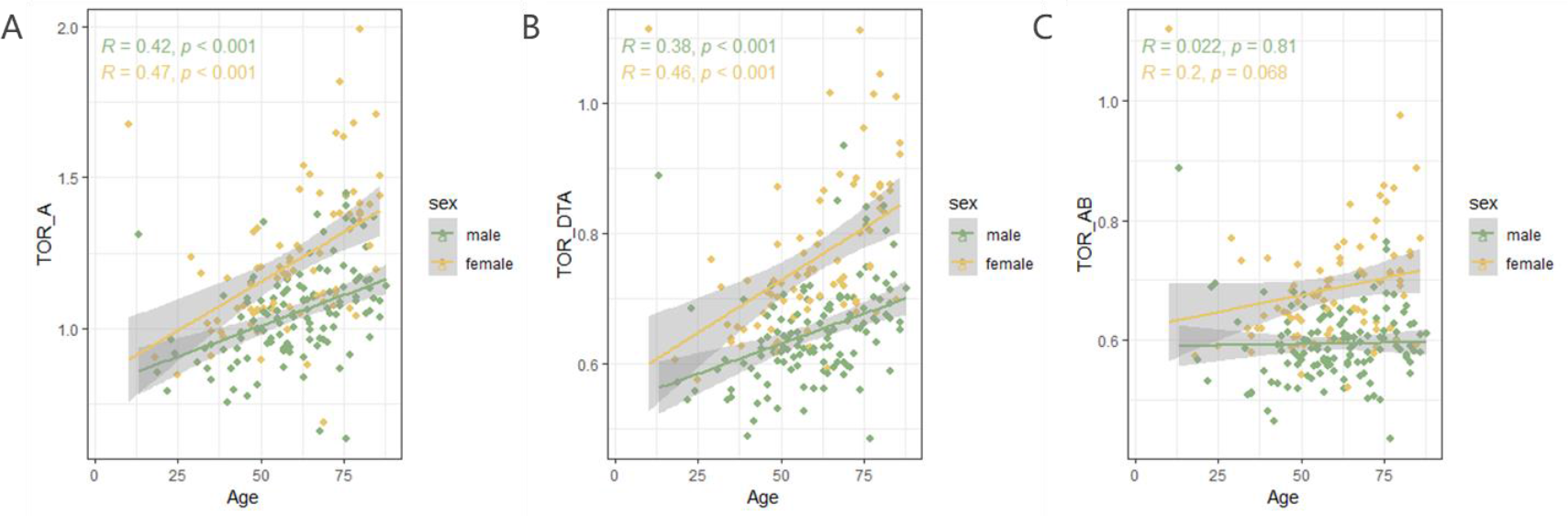
The relationship between BSA-corrected aortic tortuosity and age according to sex. Linear-regression analysis showed a linear relationship between age and aortic tortuosity(A), descending thoracic aortic tortuosity (B) in different genders. Non-linear relationship existed in abdominal aortic tortuosity either in men or women.

**Table 3.**
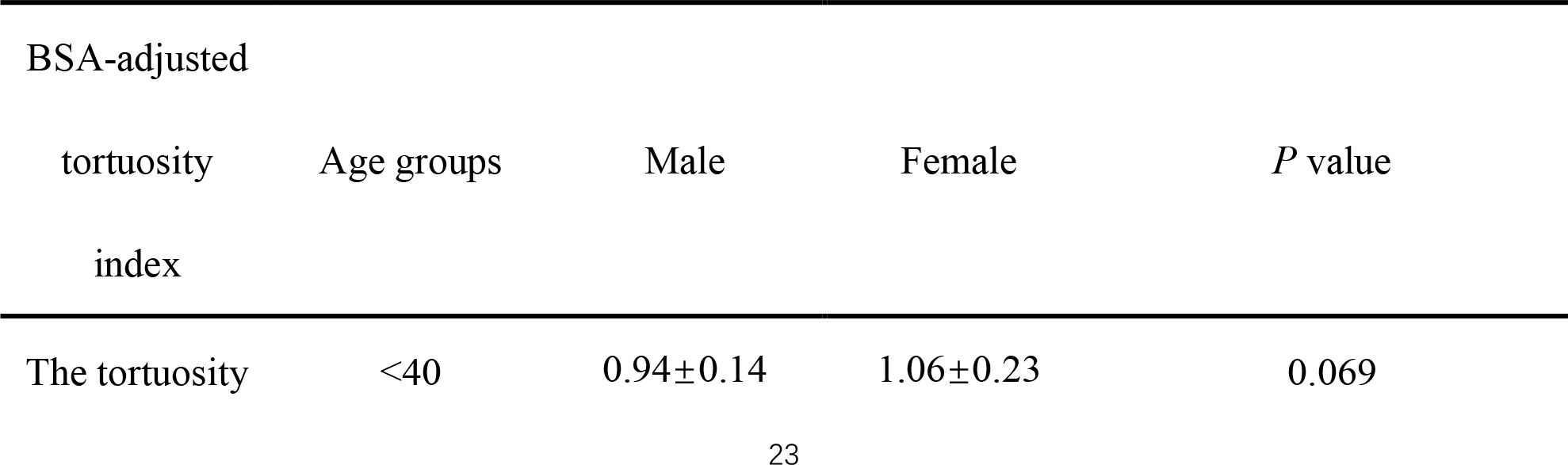
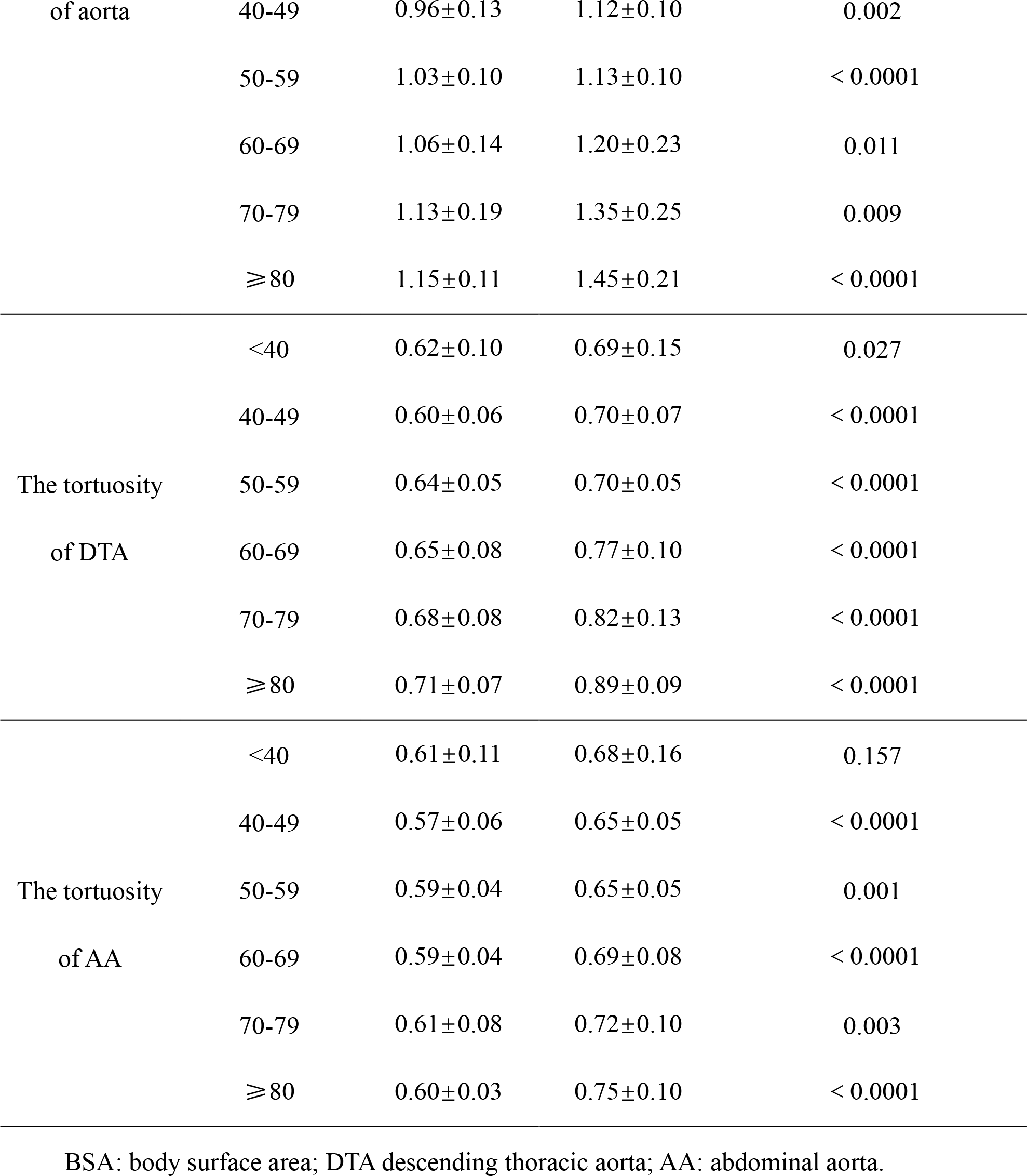
Mann-Whitney analysis of differences in disease burden between GHE disease states with or without drug approvals 2010–2019

### More remarkable age-related increase of arterial stiffness in females

We also analyzed the association between age and baPWV, which is regarded as an indicator of arterial stiffness in different sexes. The pattern was similar to that of aortic tortuosity. The study found that baPWV increases with age in both males and females, but the increase is more pronounced in females. Although males initially showed higher arterial stiffness, females eventually surpassed them as they aged. (Fig 4).

**Figure 4.**
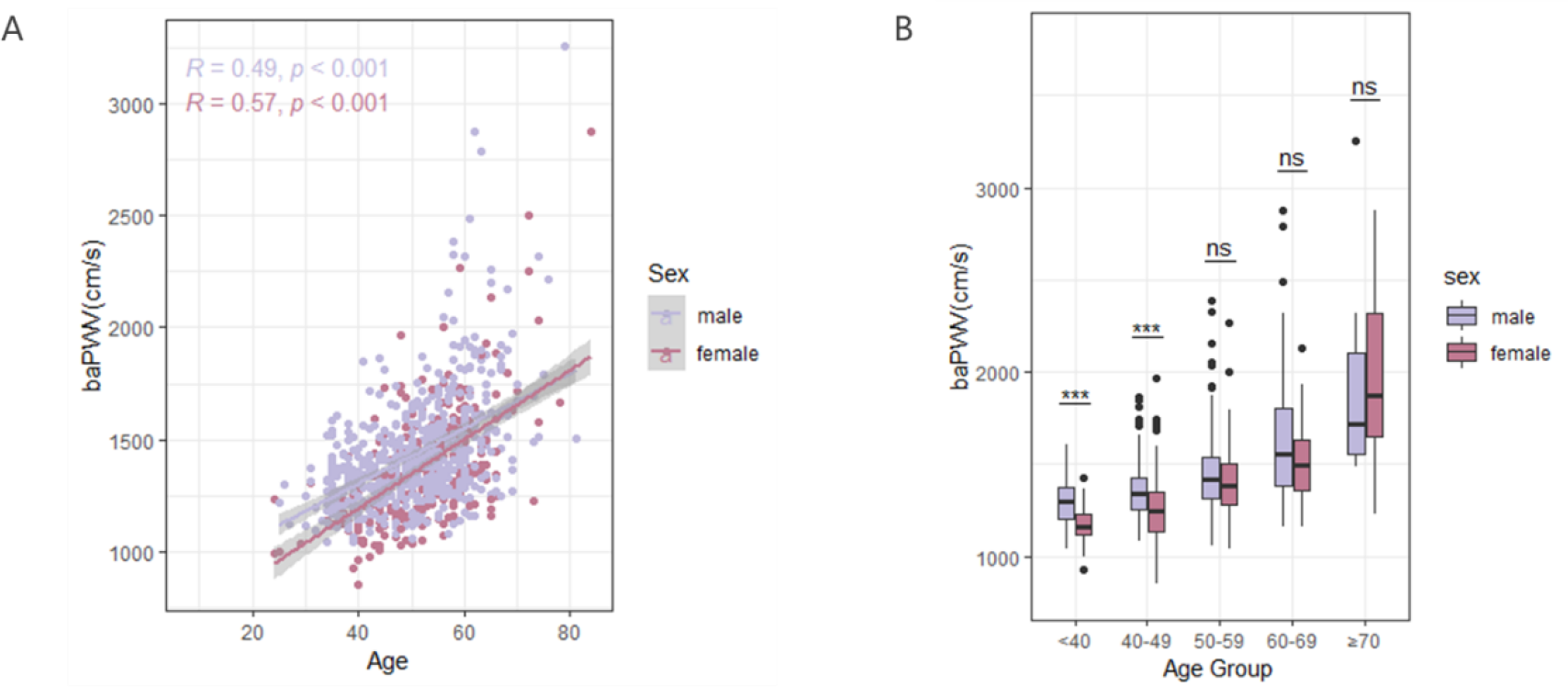
The relationship between baPWV and age according to sex (A). Sex disparities in baPWV in different age groups (B).

## Discussion

Vascular aging is an inevitable and progressive process, especially age-related changes in the arteries affect cardiovascular remodeling and the function of various organs throughout the body, as the physician Thomas Sydenham posited in the 17th century, “a man is as old as his arteries”(17). Meanwhile, the sexual dimorphism in the CVD prevalence has garnered more attention in recent years. Furthermore, investigations into sex differences in cardiovascular aging, a major risk factor for cardiovascular disease, have commenced. Our study unveiled the sexual dimorphism in age-related morphological changes of the whole aorta, in which women experience more significant arterial changes as aging compared to men, especially in terms of tortuosity. To validate our results on sex morphological differences in arteries, we also examined the age-related changes of baPWV in various sexes using a public database and found that the same intersexual difference was present in arterial elasticity.

This sex difference does not arise accidentally. Coutinho and Dart et al. demonstrated that women were more prone to developing greater pulse pressure and arterial stiffness with aging(18, 19). A study conducted by Cheng et al. showed a similar sex-specific trend in left ventricular wall thickness with aging. Men have greater absolute values of left ventricular (LV) wall thickness at all ages than women. While after adjusting for body size, women got a greater increase in LV wall thickness, especially when coexisting with other risk factors like hypertension and diabetes(20). Regarding cardiac function, the age-related increase of LV ejection fraction and diastolic dysfunction are also more remarkable in women(21, 22). The finding of anatomical aging in women in our study reflected by a more tortuous, dilated aorta and stiffness, may offer insight into sex-heterogeneity in cardiovascular function.

The sex disparity in age-related morphological changes of a particular aortic segment has garnered attention in recent years. The MESA Study indicated that the diameter of the ascending aorta increased with age. After adjustment for BSA, the normal value of females was larger than that of males in all age groups(14). Studies on the aortic arch found that aortic diameters and length were higher in men than in females, tortuosity index remained similar for both sexes(23). The results of the Rotterdam study on age-related changes in thoracic aortic diameter showed that the diameters of the ascending and descending thoracic aorta were larger in men than in women, after adjustment for body size(24). However, the aforementioned study only focused on sex differences in age-related changes in the structure of segmental arteries and did not provide a complete understanding of how age affects the structure of the entire aorta. Our findings indicate that there is no significant difference in the diameter of the abdominal aorta, ascending aorta, and descending thoracic aorta between males and females until the age of 80.

However, after 80 years of age, women have larger diameters in their abdominal aortas and lower ends of their descending thoracic aortas compared to men. In addition, after the age of 40 years old, the tortuosity of all aortic segments was significantly greater in women than in men. This suggests that the sex difference in aorta aging is predominantly manifested in tortuosity rather than diameter. Female’s arterial remodeling in the longitudinal direction is more significant than male during the lifespan. The density of smooth muscle and elasticity of elastic fibers and collagen fibers in the middle layer of the vessel wall are responsible for arterial expansion, contraction, and stress distribution, wherein reduced elastin is associated with the lengthening of the aorta(25). Therefore, sex differences in age-related changes in aortic wall microstructure should be concerned in subsequent studies. On the other hand, when combined with our analysis of the age-dependent changes in baPWV across sexes using a public database, it becomes apparent that young and middle-aged men exhibited reduced aortic tortuosity and increased arterial stiffness, resulting in greater pressure on the descending arch due to blood flow scouring, which may contribute to the higher incidence of aortic dissection. This may provide an explanation for the higher incidence of aortic dissections in males as well as the younger age at onset compared to females(26).

There is a strong correlation between age and aortic regional diameter in our study, with each part of the aorta increasing in diameter as individuals age, regardless of sex. We also noted that women experienced a more rapid expansion of their aortic diameter compared to men. Specifically, we observed that the diameter of the descending aorta to the middle segment of the abdominal aorta changed more significantly. Besides, age-specific alterations were observed in both whole aortic and descending thoracic aortic tortuosity, but not in abdominal aortic tortuosity. The results of this study indicate the importance of taking regional variations into account while studying aortic ageing. Additionally, it suggests that when evaluating vascular ageing, it may be more effective to focus on the morphology of the descending aorta.

Although our observational study didn’t demonstrate the specific mechanism by which sex directly influences the geometry of aortic aging, it is possible to argue from the results that estrogen plays an important role(27). In general, estrogen exerts a protective effect on vascular wall in premenopausal women. Estrogen protects the vascular wall in premenopausal women with receptors highly expressed in endothelial and vascular smooth muscle cells throughout the human body(28). The mechanism behind estrogen-induced vasorelaxation is believed to involve the interaction of estrogen with the vascular smooth muscle, endothelial cells, and the vessel wall(29, 30). Relevant research has found that estrogen can exert cardiovascular protective effects by inhibiting the intravascular accumulation of collagen and the proliferation of vascular muscle cells (VSMCs)(27). Our study reveals a more significant age-related change in the aorta among women, particularly middle-aged and older women, suggesting an innate effect of sex on aortic aging. The decline in estrogen levels after menopause mat accelerate this process. More research is warranted to elucidate the concrete mechanisms.

The findings of this study should be interpreted with caution due to several limitations.

Firstly, the data was extracted from a small-scale observational cohort study conducted at a single center, which may limit the generalizability of the results. Additionally, confounding factors may have influenced the outcomes. Secondly, non-ECG gating contrast CT was used to measure aortic diameters and tortuosity, which did not consider the impact of contraction and diastole on aortic morphologies. As a result, the accuracy of the measurements may be limited. Inevitably, some of our subjects had malignant tumors which may affect the morphology of the aorta. To validate our findings and illustrate the underlying mechanisms, it is imperative to conduct larger-scale surveys with multi-center involvement and high-precision measurements of the normal population.

## Conclusion

Patterns of vascular aging in aortic morphology differ between the sexes across the life course, with females experiencing more significant changes, particularly in advanced age groups. This implies that female arteries are more vulnerable to aging. Therefore, it is crucial to prioritize effective prevention strategies for vascular aging and related diseases, focusing on females who face greater disparities and stand to benefit most from preventive measures.

## Data Availability

The datasets analyzed during the current study are available from the corresponding author on reasonable request.

## Acknowledgements

Zhijie Jian and Yue Wu conceived and designed the research; Lele Cheng and Zixuan Meng analyzed and interpreted the patient data; Guolin Yao and Jian Yang was responsible for the quality control of the CT scans; Hui Liu and Yue Yu collected clinical data; Lele Cheng and Zixuan Meng performed statistically analysis and drafting of the manuscript; Wenjun Liu, Zhijie Jian, Yue Yu and Yue Wu revised manuscript; Zhijie Jian and Yue Wu had primary responsibility for final content. All authors read and approved the final manuscript.

## Ethics Statement

The studies involving human participants were reviewed and approved by the Ethics Committee approval of the First Affiliated Hospital of Xi’an Jiaotong University. The patients/participants provided their written informed consent to participate in this study.

